# The Effect of Area Deprivation on COVID-19 Risk in Louisiana

**DOI:** 10.1101/2020.08.24.20180893

**Authors:** KC Madhav, Evrim Oral, Susanne Straif-Bourgeois, Ariane L. Rung, Edward S. Peters

**Affiliations:** Epidemiology Program, School of Public Health, Louisiana State University Health Sciences Center New Orleans, New Orleans, LA, United States; Biostatistics Program, School of Public Health, Louisiana State University Health Sciences Center New Orleans, New Orleans, LA, United States

## Abstract

**Purpose:** Louisiana currently has the highest per capita case count for COVID-19 in the United States and disproportionately affects the Black or African American population. Neighborhood deprivation has been observed to be associated with poorer health outcomes. The purpose of this study was to examine the relationship between neighborhood deprivation and COVID-19 in Louisiana.

**Methods:** The Area Deprivation Index (ADI) was calculated and used to classify neighborhood deprivation at the census tract level. A total of 17 US census variables were used to calculate the ADI for each of the 1148 census tracts in Louisiana. The data were extracted from the American Community Survey (ACS) 2018. The neighborhoods were categorized into quintiles as well as low and high deprivation. The publicly available COVID-19 cumulative case counts by census tract was obtained from the Louisiana Department of Health website on July 31, 2020. Descriptive and Poisson regression analyses were performed.

**Results:** Neighborhoods in Louisiana were substantially different with respect to deprivation. The ADI ranged from 136.00 for the most deprived neighborhood and -33.87 in the least deprived neighborhood. We observed that individuals residing in the most deprived neighborhoods had a 45% higher risk of COVID-19 disease compared to those residing in the least deprived neighborhoods.

**Conclusion:** While the majority of previous studies were focused on very limited socio-environmental factors such as crowding and income, this study used a composite area-based deprivation index to examine the role of neighborhood environment on COVID-19. We observed a positive relationship between neighborhood deprivation and COVID-19 risk in Louisiana. The study findings can be utilized to promote public health preventions measures besides social distancing, wearing a mask while in public and frequent handwashing in vulnerable neighborhoods with greater deprivation.

## 1. Introduction

On March 09, 2020, Louisiana reported its first case of COVID-19 and soon thereafter appeared to be a hot spot of the coronavirus pandemic in the US [1]. Within two weeks of the initial confirmed case, the state had one of the world’s highest average daily growth rate [2-4]. As of July 31, 2020, the state of Louisiana has the highest per capita case count in the United States with a total of 116,280 confirmed cases and 3,835 deaths [2]. The incidence and mortality rates of COVID-19 has been disproportionate across racial and ethnic groups [5, 6]. Specifically, non-Hispanic African Americans have higher rates of incidence, hospitalization, and death from COVID-19 compared to non-Hispanic Whites. In early July, the US Centers for Disease Control and Prevention (CDC) estimated that non-Hispanic African Americans have 4.7 times the rate of age-adjusted COVID-19 related hospitalization rates than non-Hispanic Whites [7]. The sources of disparities in COVID-19 outcomes might be explained from a social determinants of health perspective. Non-Hispanic African Americans are more likely to have vulnerable and low-paying jobs that don’t allow remote work, which increases risk of contracting COVID-19 [8, 9]. Furthermore, non-Hispanic African Americans are more likely to rely on public transportation and to live in crowded housing or work in crowded worksite that places them an increased risk for COVID-19 disease. African Americans exhibit a greater burden of chronic medical conditions, such as hypertension, diabetes, heart disease, chronic disease, and obesity that increase the severity of COVID-19 illness [10-12]. In Louisiana, 2.9 million people have at least one chronic condition, and a total of 68 percentage of Louisiana adults are overweight or obese [13]. Furthermore, the poverty rate is much higher among African Americans as those of non-Hispanic Whites, and are concentrated in neighborhoods with high poverty [14, 15]. The neighborhood socioeconomic status (SES) is linked to access to health care services, people residing in low SES neighborhoods are less likely to have access to health care services, which further increases the risk of adverse health outcomes related to COVID-19, such as higher hospitalizations and mortality [16, 17].

Preliminary reports show a relationship between the neighborhood of residence and COVID-19 disease, hospitalization and death [18-20]. Those who reside in deprived neighborhoods, defined by low income and education, higher unemployment, and substandard living conditions have a greater risk of poor health outcomes such as obesity, diabetes, cancer, and heart diseases [21-23]. Higher incidence and mortality from COVID-19 have been observed in low-income or deprived neighborhoods [24-26]. A study conducted by Bilal et al. reported a 36% higher incidence of COVID-19 in deprived neighborhoods compared to less deprived neighborhoods [25]. Systemic health, social, and income inequities are considered as the primary reasons that have contributed to the increased risk of contracting COVID-19 in persons residing in deprived neighborhoods [7, 27, 28].

Risk factors leading to COVID-19 disease, hospitalization, and mortality are not only at the individual or biological level; neighborhood-level factors and their interactions with individual-level factors are also responsible for the observed disparities. Lack of access to health care, unemployment, less education, and poor housing conditions significantly increase the risk of COVID-19 infection [28-31]. These determinants of health can be studied collectively as neighborhood or area deprivation.

Socioeconomic characteristics of residential neighborhoods influence health-related behaviors, conditions, and health outcomes [32, 33]. Deprived neighborhoods are correlated with health risk behaviors, overcrowding, less social cohesion, and higher levels of environmental pollutants, and has been identified as a critical social determinant of health [34-37]. Low socioeconomic status (SES), often regarded as a fundamental cause of disease, has been shown to increase the risk of COVID-19 because it impacts access to fundamental resources that an individual or a neighborhood may require to avoid COVID-19 [24, 38].

Neighborhoods with a higher number of people per household or room tend to have a higher rate of confirmed COVID-19 cases than neighborhoods with fewer residents [25, 39, 40]. Individuals who share a room or live in overcrowded housing and the use of public transportation often spread the disease rapidly as distancing preventive measures are impossible to adopt.

In this study, we used the Area Deprivation Index (ADI) to measure neighborhood deprivation. The ADI is a composite measure of neighborhood socioeconomic disadvantage, created by Gopal K Singh in 2003 [41]. The ADI, composed of 17 education, employment, housing-quality, and poverty census derived measures, is a robust metric measuring many relevant social determinants of health that may help explain the socio-biologic mechanisms of disease [41, 42]. We hypothesize that deprived Louisiana neighborhoods have a higher risk of COVID-19 reported cases than less deprived neighborhoods. To date, few studies in the US and none in Louisiana have assessed the role of social determinants of health on COVID-19 disease. The studies that exist are limited, examining only a couple of specific risk factors, such as overcrowding and income. The use of the Area Deprivation Index (ADI) in the present study includes 17 neighborhood-level factors and provides a robust measure of neighborhood deprivation. The primary purpose of this paper is to investigate the relationship between neighborhood deprivation and COVID-19 risk in Louisiana.

## 2. Materials and methods

### 2.1 Study Data

Publicly available data on cumulative COVID-19 cases by census tract was obtained from the Louisiana Department of Health website on July 31, 2020 [2]. There are 64 parishes (counties) and 1,148 census tracts in Louisiana. All 64 parishes have reported cases of COVID-19. Because the census tract is considered a good proxy for neighborhood, census tract was selected as the unit of analysis for this study.[43] We extracted the American Community Survey (ACS) 2018 data for census tract level measures for Louisiana [44].

### 2.2 COVID-19

The main outcome in this study was COVID-19 cases per 1,000 persons in Louisiana census tracts as of July 31, 2020.

### 2.3 Neighborhood Deprivation

Neighborhood deprivation was measured by the ADI, as described by Singh in 2003 [41]. ADI is a validated, factor-based deprivation index that uses 17 census derived measures of poverty, education, housing, and employment indicators at the census tract level to classify the neighborhoods [41, 45]. More-disadvantaged neighborhoods are those with a higher ADI score.

The census indicators used in the calculation of ADI include educational distribution (percentage of the population with less than 9 years and with 12 or more years of education), median family income, median home value, median gross rent, median monthly mortgage, income disparity, unemployment, percent employed person in white-collar occupation, percent families below poverty, percent population below 150% poverty threshold, single-parent household rate, homeownership rate, percent household without a telephone, percent household without a motor vehicle, percent occupied housing units without complete plumbing, and household crowding [41, 45].

#### Calculation of ADI score

Data from the Census Bureau’s American Community Survey (ACS) 2018 was used to calculate the ADI score. The 17 US census indicators were multiplied by the Singh’s coefficients (factor weights) for all census tracts in Louisiana [41, 46].

The base score of each indicator was summed to get the total base score for a census tract. Each census tract’s base score was standardized by dividing the difference between the individual census tract base score (*b*) and the Louisiana census tract population mean (p), by Louisiana census tract population standard deviation (*S_p_*) [46].

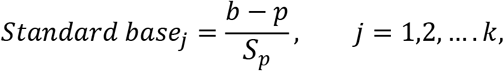

where *j* represents the *j^th^* census tract, and *k* is the total number of census tracts in Louisiana. Finally, the standardized values were adjusted to a base mean of 100 and a standard deviation of 20 as suggested by Knighton et al [46].

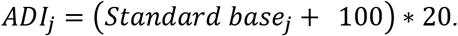

The details of ADI calculation and a list of variables included in the calculations can be found in Knighton et al [46].

Based on the ADI scores, the census tracts were categorized into quintiles of deprivation; they were also dichotomized as either low deprivation or high deprivation. The median Louisiana ADI was used to dichotomize the census tracts. Census tracts with missing values for the indicators were excluding while calculating the ADI.

### 2.3 Statistical Analysis

SAS 9.4 software was used for statistical analyses. Heat maps were created using ArcGIS software. Mean, standard deviation, median and interquartile range (IQR) of census indicators by ADI Quintiles (least deprived: Q1 and most deprived: Q5) were calculated for all census tracts in Louisiana. Poisson regression analysis was performed to estimate risk ratio. An offset variable was used, and the model was corrected for over dispersion.

## 3. Results

There was a substantial difference between the ADI of the least deprived and most deprived neighborhoods. The overall median (IQR) ADI for Louisiana was 104.32 (76.00), with the most deprived neighborhood having an ADI of 136.00, and the least deprived neighborhood having an ADI of -33.87. While the median ADI of the least deprived neighborhood was 76.00, the median ADI of the most deprived neighborhood was 118.45 (Table 1).

**Table 1:** Area Deprivation Index (ADI) distribution in Louisiana Census Tracts (N=1127)

|  | **LA**<br>**(Overall)** | Q1<br>(Least<br>Deprived) | Q2 | Q3 | Q4 | Q5<br>(Most<br>Deprived) |
| --- | --- | --- | --- | --- | --- | --- |
| Mean | **100** | 69.29 | 96.25 | 104.26 | 110.85 | 119.30 |
| Std Dev. | **20.00** | 20.09 | 3.03 | 1.98 | 1.91 | 4.61 |
| Median | **104.32** | 76.00 | 96.64 | 104.32 | 110.94 | 118.48 |
| IQR | **18.82** | 20.09 | 5.02 | 3.50 | 3.46 | 5.37 |
| Minimum | **-33.87** | -33.87 | 89.82 | 100.88 | 107.65 | 114.01 |
| Maximum | **136.00** | 89.61 | 100.86 | 107.62 | 114.00 | 136.00 |

Table 2 shows the median and interquartile range of census indicators that were used in the calculation of ADI. The most deprived neighborhoods in Louisiana had 31.02% of families below poverty. Similarly, more than 15.47% of occupied housing units lacked a motor vehicle. The unemployment rate was more than twice as high in the deprived neighborhoods as the less deprived neighborhoods. Almost 3% of households in the most deprived neighborhoods had more than one person per room. Similarly, the median home value in the most deprived neighborhood was substantially lower than those in the least deprived neighborhoods ($74,550 vs $273,900). These results suggest that poor people with lower levels of education were clustered together in Louisiana.

**Table 2:** Median and IQR values of census tract level indicators in Louisiana.

| Indicators | Least Deprived<br>Neighborhoods<br>(Q1) |  | Most Deprived<br>Neighborhoods<br>(Q5) |  |
| --- | --- | --- | --- | --- |
|  | Median | IQR | Median | IQR |
| Percent of population aged ≥ 25 years with < 9 years of education (%) | 1.71 | 2.29 | 6.68 | 5.97 |
| Percent of population aged ≥ 25 years with greater or equal to a high school (%) | 94.57 | 5.40 | 75.48 | 10.50 |
| Percent of employed person ≥16 years of age in white-collar occupations (%) | 49.79 | 14.50 | 20.13 | 10.63 |
| Median family income (\$) | 96,071 | 29,940 | 32,410 | 11,959 |
| Income disparity* | 0.83 | 0.49 | 1.82 | 0.47 |
| Median home value (\$) | 273,900 | 112,600 | 74,550 | 21,750 |
| Median gross rent (\$) | 1106 | 280 | 687 | 203 |
| Median monthly mortgage (\$) | 1803 | 575 | 881 | 171 |
| Percent of owner-occupied housing units (%) | 71.19 | 30.11 | 47.18 | 28.01 |
| Percent of civilian labor force population ≥ 16 years of age unemployed (%) | 4.09 | 3.36 | 10.61 | 9.03 |
| Percent of families below the poverty level (%) | 4.95 | 5.71 | 31.02 | 16.70 |
| Percent of the population below 150% of the poverty threshold (%) | 14.44 | 12.07 | 53.15 | 15.06 |
| Percent of single-parent households with children < 18 years of age (%) | 6.94 | 5.87 | 20.33 | 11.64 |
| Percent of occupied housing units without a motor vehicle (%) | 4.11 | 6.53 | 15.47 | 13.78 |
| Percent of occupied housing units without a telephone (%) | 1.50 | 1.60 | 3.25 | 3.15 |
| Percent of occupied housing units without complete plumbing (%) | 0.00 | 0.00 | 0.00 | 0.92 |
| Percent of occupied housing units with more than one person per room (%) | 0.62 | 1.48 | 2.87 | 3.88 |

*\*Income disparity was defined as the log of 100\*ratio of the number of households with* *<\$10,000 income to the number of households with \$50,000+ income*

From the Poisson regression analysis (Table 3) we observed that census tracts with high neighborhood deprivation was associated with an increased risk of COVID-19 in Louisiana. There was a 45% higher risk of COVID-19 disease among individuals residing in most deprived neighborhoods compared to those in the least deprived neighborhoods (RR=1.45, 95% CI=1.31-1.59).

**Table 3:** Relationship between the ADI and COVID-19 in Louisiana census tracts (N=1127)

| ADI | N | RR | 95% CI | p-value |
| --- | --- | --- | --- | --- |
| Low Deprived Neighborhoods | 563 | Reference | - | - |
| High Deprived Neighborhoods | 564 | 1.45 | (1.31-1.59) | <0.0001 |

In figure 1, the census tracts in red represent the most deprived neighborhoods, while the census tracts in green are the least deprived neighborhoods in Louisiana. In figure 2, the census tracts in yellow represent census tracts with fewer COVID-19 cases per 1,000 persons as of July 31, 2020, while the census tracts in brown and dark brown represent higher COVID-19 cases per 1,000 persons. Figure 3 shows the distribution of ADI and COVID-19 cases per 1,000 persons simultaneously in Louisiana by census tracts.

**Figure 1:**
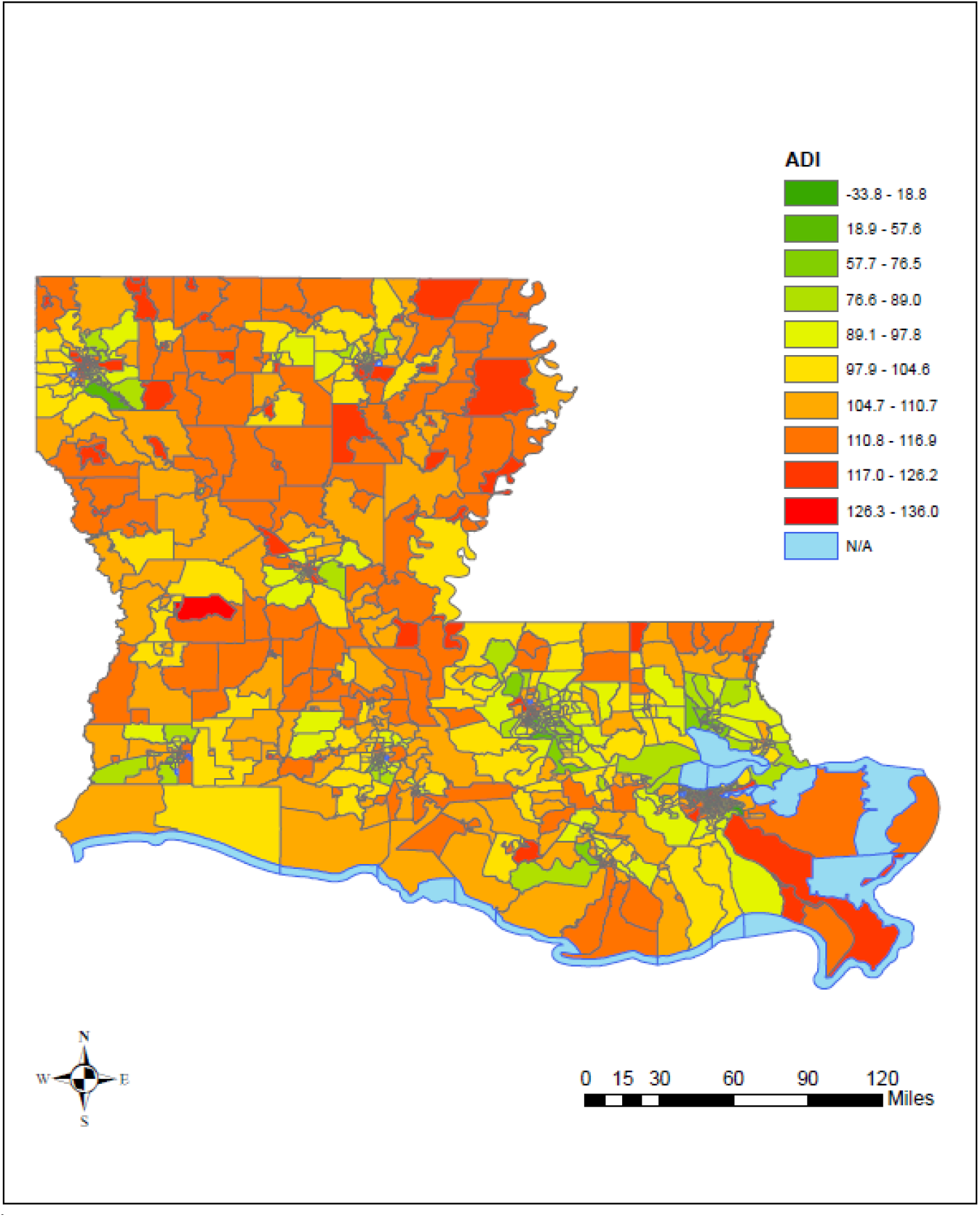
ADI in Louisiana census tracts

**Figure 2:**
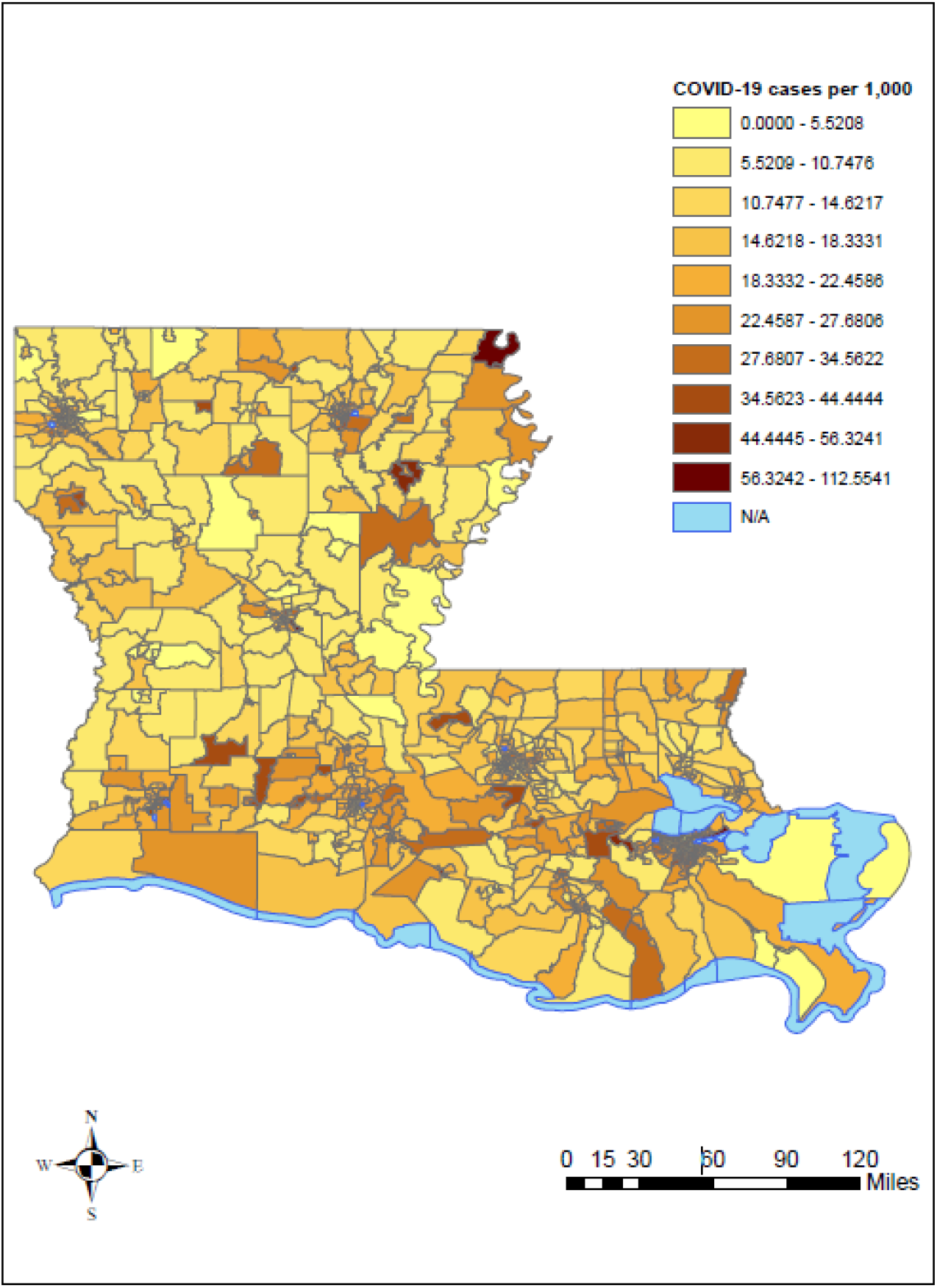
COVID-19 cases per 1,000 persons in Louisiana by census tract

**Figure 3:**
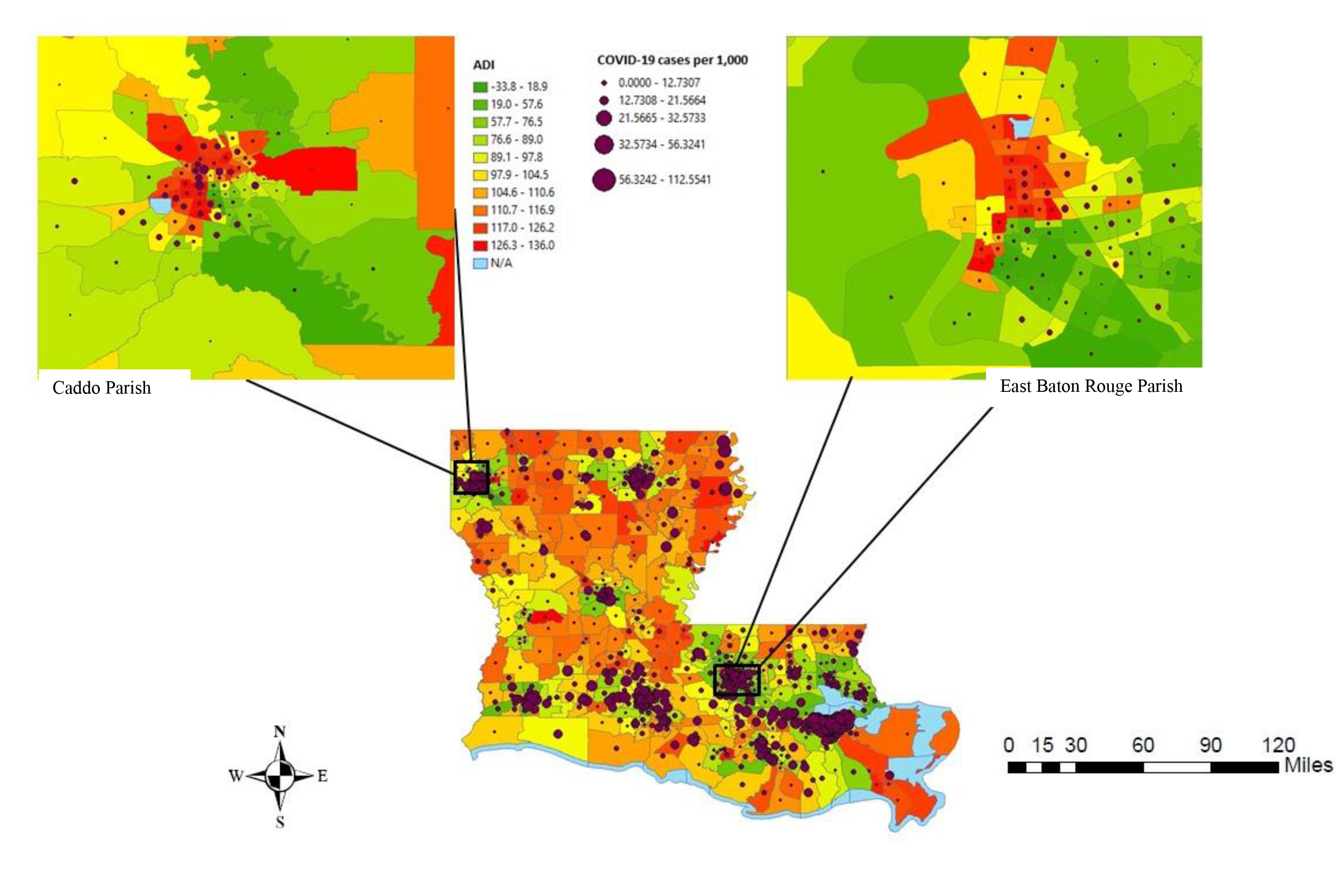
ADI and COVID-19 cases per 1,000 persons in Louisiana by census tracts

## 4. DISCUSSION

To our knowledge, this is the first study to investigate the role of neighborhood deprivation on COVID-19 in Louisiana. While previous studies were limited to a very few socio-environmental factors, we used a composite area-based deprivation index to identify neighborhood deprivation in Louisiana, US. The ADI includes 17 US census indicators and could serve as an important tool in assessing the role of the neighborhood on COVID-19 disease. Our findings demonstrated the increased rate of COVID-19 disease among individuals who live in the most deprived neighborhoods compared to individuals residing in the least deprived neighborhoods. A New York study showed higher infection rates in low-income communities in New York City [47, 48]. The neighborhood or built environment can impact health status either by influencing environmental quality or by influencing behaviors that impact the transmission of COVID-19. One of the major factors that might have fueled the spread of COVID-19 disease in poor neighborhoods is likely to be overcrowded living spaces. A study conducted by Emeruwa et al. observed a strong association between neighborhood socioeconomic status and household crowding and COVID-19 cases in New York City [48]. The odds of infection were twice as high among individuals who lived in households with greater crowding (interdecile OR, 2.27 [95% CI, 1.12-4.61]). Similarly, a study in California showed 3.7 times the rate of confirmed COVID-19 cases in overcrowded neighborhoods compared to less crowded neighborhoods.[49] These findings illustrate how the housing environment plays an important role in disease dynamics and in determining the health of individuals. Neighborhood socioeconomic status and overcrowded housing may explain why non-Hispanic African American and Hispanic populations are at higher risk of getting COVID-19.

In addition to overcrowding and neighborhood-level SES, the disparities in COVID-19 cases between neighborhoods might be directly related to the nature of residents’ occupations, a lack of telecommunication infrastructure, use of public transportation, and utility disruptions. Areas with concentrated poverty and extreme racial segregation had a higher incidence of COVID-19 [5, 50, 51]. Since low-income individuals often lack private vehicles, and may rely on public transportation, this in turn increases the risk of contracting COVID-19. An early study in China observed a positive association between the frequency of public transportation use and cumulative cases of COVID-19 [52]. However, the results may not be generalizable to areas where public transportation is not available, especially in rural Louisiana. In New York, Carrion et al. also found higher subway ridership among individuals who reside in neighborhoods where COVID-19 cases were higher [24]. Another risk factor that could potentially increase the individual’s risk of contracting infection is occupation. Although many individuals have been practicing social distancing by working from home, 71% of American workers cannot work from home [53]. Individuals in certain blue-collar jobs tend to have a higher incidence of and mortality from COVID-19 [54]. The symptomatic cases of COVID-19 are easily picked up and can help prevent the spread of disease by isolating themselves, however, asymptomatic cases and symptomatic individuals who don’t get paid sick leave or are essential workers are likely to spread the disease more rapidly. As of July 29, 2020, there were more than 100 worksite outbreaks in Louisiana [2]. The majority of studies have emphasized how adversely affected by COVID-19 certain racial and ethnic communities are; however, these groups of people may have differential exposure to the virus due to long-standing systemic health and social inequalities.

This study has several limitations. Due to a lack of data, we were unable to account for COVID-19 testing per census tract in our statistical analysis or perform a time series analysis of COVID-19 case counts. Similarly, we couldn’t include the data on COVID-19 testing in a deprived neighborhood and the availability of free testing clinics in our analysis. This study is limited only to COVID-19 cases per 1,000 persons in Louisiana census tracts, the severe outcomes such as hospitalizations including Intensive Care Units (ICUs) admissions, and mortality were not assessed. Another limitation is that the impact of race couldn’t be examined due to the lack of data at the census tract by race.

A key strength of this study is the use of the ADI to characterize neighborhood disadvantage. The ADI is a validated and becoming more widely used composite index of neighborhood disadvantage. The ADI provides a robust method to identify and classify deprived neighborhoods. The use of the most relevant social determinants of health in the calculation of ADI allows for better contextualization of the neighborhood.

Despite these limitations, we believe that this study contributes to the literature on social determinants of health and COVID-19 disease in the neighborhood by establishing the relationship between the neighborhood deprivation and COVID-19 cases in Louisiana. Findings may help authorities to prioritize the public health response especially by increasing free testing sites and contact tracing in the targeted areas. In addition, it is important to promote public health preventions measures for case isolation and quarantine of close contacts besides social distancing, wearing a mask while in public and frequent handwashing to ultimately reduce the spread of COVID-19 in the most vulnerable populations.

## 5. Conclusion

We observed a great disparity in deprivation among Louisiana neighborhoods. We also found an association between neighborhood deprivation and the COVID-19 cases per 1,000 persons in Louisiana. There have been many studies on how COVID-19 is clustered in neighborhoods, however, future studies should explore specific mechanisms behind this association.

## Data Availability

Data use in this study is publicly available.

